# Delirium and Increased Risk of Developing Dementia: An Emulated Target Trial Analysis

**DOI:** 10.64898/2026.05.11.26352925

**Authors:** Cara S. Rathmell, Haoqi Sun, Wendong Ge, Colin Magdamo, Sudeshna Das, Lidia M.V.R. Moura, Sahar Zafar, Oluwaseun Akeju, Shibani Mukerji, Kendrick Shaw, M.Brandon Westover

## Abstract

**Background:** Multiple studies suggest bidirectional links between delirium and Alzheimer’s Disease and Related Dementias (ADRD). Although they establish a strong association between delirium and subsequent ADRD, it has not been explored using statistical causal inference which makes the best use of observational data to minimize biases.

**Methods:** We conducted an emulated clinical trial to estimate the effect of experiencing delirium during hospitalization between April 2017 and September 2019 on the cumulative incidence of ADRD over two years following hospital admission in patients 65 and older. The emulated trial used observational data from individuals in the Mass General Brigham Electronic Medical Record (EMR). We carried out statistical causal survival analysis using methods that adjust for confounding, censoring, competing risks, and immortal-time bias, including inverse propensity weighting (IPW) and g-formula approaches.

**Results:** Of the 6029 patients hospitalized in this time frame who were 65 or older with evidence of a PCP in the EMR, 5901 were included in the analysis based on no history of dementia diagnosis or medications 12 months prior to admission. At two years post-admission, the adjusted cumulative incidence of ADRD in individuals who did not experience delirium was 7.6% (95% Confidence Interval [CI] 4.0-12.1%) while it was 20.2% (95% CI 13.2-27.9%) for those who did experience delirium when calculated using the IPW method.

**Conclusions:** Our emulated trial results argue for a strong association between delirium during hospitalization and the risk of developing ADRD in the two years following hospital admission in individuals 65 and older.

**Key Points:** *Question:* We sought to answer whether statistical causal inference would show the same association between delirium and the onset of dementia in the two years following hospitalization.

*Findings:* Our emulated trial results argue for a strong association between delirium during hospitalization and the risk of developing ADRD in the two years following hospital admission in individuals 65 and older.

*Meaning:* The implications of demonstrating this relationship underscore the importance of delirium-mitigating interventions for long-term cognitive outcomes.

## INTRODUCTION

Multiple studies have suggested bidirectional links between delirium and Alzheimer’s Disease and Related Dementias (ADRD)^1-6^. This raises the possibility that preventing delirium might forestall the onset of ADRD^7^. However, although prior studies establish a strong association between delirium and subsequent ADRD^2^, it is unclear whether this relationship can be confirmed using statistical causal inference. Evidence for a strong association needs to come from methods that reduce confounding in observational data, emulating a randomized clinical trial^8^. Such evidence would strengthen the case for future controlled trials.

Causal inference enables emulating a randomized clinical trial. With this approach, individuals from electronic medical record (EMR) data are “randomized” into cohorts and treated as if they were participating in a trial. While this procedure has certain limitations, it offers another opportunity to demonstrate a relationship between delirium and dementia, with the added benefit of controlling for biases that other observational study designs do not.

Past observational studies have not accounted for immortal time bias, a bias introduced by allowing individuals to be deemed at risk of an outcome when they are not. By introducing the emulated clinical trial, we control for this bias and thus further underscore the strength of the relationship between delirium and ADRD.

In this study, we used a large-scale EMR dataset to investigate the effect of delirium on the risk of developing ADRD. Specifically, we emulate a clinical trial using observational data to study the effect of an individual experiencing delirium during hospitalization on their risk of developing ADRD over the two years following hospitalization.

## METHODS

We used an emulated target trial approach to emulate a hypothetical randomized clinical trial^8,9^. Specifying the ideal randomized trial to answer the research question forces a rigorous formulation of the study design components and assumptions necessary to emulate the trial using observational data^8^.

We consider a target trial in which patients are randomly assigned to a hypothetical “delirium” subset or not; in effect, we assume complete experimental control over whether patients develop delirium. In practice, multiple exposures can increase the risk of delirium, including administration of benzodiazepine drugs, hospital schedules (e.g., exams, blood draws) that disrupt sleep, and existing interventions that prevent delirium, most notably the Hospital Elder’s Life Program (HELP)^6,10-12^ and dexmedetomidine^13-15^, are not 100% effective. Nevertheless, for our hypothetical target trial, we assume full experimental control to focus on the question: What is the effect of delirium on the development of Alzheimer’s disease or related dementias? By answering this question, we aim to provide an upper bound on the effects that real-world delirium-prevention interventions could have on incident dementia risk.

## Target trial specification

Table 1 summarizes the key protocol components. The target trial to answer the question of interest would randomly assign eligible patients at the time of hospital admission to one of the 2 treatment strategies: (1) develop delirium within 14 days of hospital admission, or (2) do not develop delirium within the same 14-day period. The 14-day period was chosen to be long enough that most delirium incidents will fall within this window. Patients who develop delirium within this period are considered adherent to the assigned exposure. The outcome is dementia (Alzheimer’s disease and related dementias, ADRD), evaluated in a follow-up period of 2 years following treatment. Trial eligibility criteria include age 65 or greater, community-dwelling, and no history of ADRD over the prior 12 months before hospitalization. The following sections describe the observational study that we designed to emulate this target trial.

**Table 1.** Target Trial vs Emulated Trial.

|  | Target Trial | Emulated Trial |
| --- | --- | --- |
| Eligibility Criteria | Hospitalized at MGB April 1, 2017 to September 1, 2019<br>Age $\geq 65$ years<br>Community dwelling/PCP in EHR<br>No history of ADRD as reported by patient/family on day of admission<br>No history of ADRD medication use | Same<br>Same<br>Evidence of PCP visit in EHR in previous 12 months<br>No history of ADRD prior to admission the 12 months prior to admission as determined by NLP algorithms<br>No documented prescription of ADRD medications for previous 12 months |
| Exposure ("Treatment" strategies) | "Treatment" arm: "Delirium", as defined by CAM, within the first 14 days after admission<br>Control arm: "No delirium" within the first 14 days after admission | "Treatment" arm: "Delirium", as defined by NLP algorithm, within the first 14 days after admission<br>Control arm: "No delirium" within the first 14 days after admission |
| Assignment | Randomly assign "Delirium" or "No delirium" within 14 days of enrollment at time of hospitalization | Emulated randomization by balancing baseline confounders using IPTW* for treatment selection |
| Outcome | Time to ADRD from admission | Same. Time to ADRD detected in EHR data based on ICD Codes and Medications |
| Follow-up | Starts at randomization (at admission)<br>Ends at date of ADRD diagnosis, | Starts at admission<br>Ends at date of ADRD diagnosis, death, loss to follow-up (date of last use of |
|  | death, loss to follow-up, or end of study (i.e., 2 years post hospital admission); whichever occurs first | EHR), or end 2 years post hospital admission; whichever occurs first |
| Causal Contrast | Per-protocol effect** | Observational analogue of per-protocol effect** |
| Statistical Analysis | Survival analysis of time to ADRD, accounting for censoring with semi-competing risk | Causal survival analysis of time to ADRD, accounting for censoring with semi-competing risk and baseline confounding |

Legend Table: MGB=Mass General Brigham; PCP=Primary Care Provider; EHR=Electronic Health Record; NLP=Natural Language Processing; IPTW=Inverse Probability of Treatment Weights; ICD=International Classification of Diseases; IPCW=Inverse Probability of Censoring Weights.
\*Observational analog of intention-to-treat analysis may involve using Cox Proportional Hazards regression with IPTW and IPCW.
\*\*Analysis completed is consistent with “intention-to-treat” due to lack of treatment adherence weighting, but the effect can be considered “per-protocol” in practice due to the lack of patients who did not “adhere” to their assignment.

### Per protocol statistical analysis of the target trial

We aim to estimate the effect of delirium under full adherence to the protocol of the target trial: the per-protocol effect^16,17^. Specifically, we are interested in the difference between the cumulative incidence of ADRD between treatment groups. To estimate this per-protocol effect in the target trial, we would use a time-varying hazards model, approximated via a pooled logistic regression model^18-20^ that includes an indicator for treatment group assignment, a flexible function of time (e.g., a second-order polynomial), and the baseline covariates. Our analysis approach would censor patients if they deviated from their assigned treatment strategy because of failure to adhere to the assigned treatment, loss to follow-up, or death. A patient’s data is included in the estimation of the initial part of the cumulative incidence curve for the assigned group, but it ceases to contribute after the time of “non-adherence” (developing delirium in the “no delirium” group).

To adjust for the potential selection bias due to censoring^16,21,22^, each patient would receive an inverse probability weight^9^. The denominator of these weights is the individual’s probability of adhering to the assigned treatment, conditional on baseline prognostic factors (see below), which in turn is the product of three probabilities of being censored due to non-adherence, loss to follow-up, or death. Truncating these weights at the 99.5th percentile would reduce the effects of outliers on our results.

An important assumption is that the adjustment includes all important baseline variables that influence the risk of the primary outcome and that predict adherence to the assigned treatment. In our target trial, we adjust for patient attributes and comorbidities relevant to the risk for developing delirium and dementia; these include age, sex, race, and burden of chronic illness,which we would measure using the Charlson Comorbidity Index (CCI)^2,23-26^. When race was missing, we categorized those patients as “not specified”. We don’t adjust for complications that develop after trial enrollment, such as metabolic derangements or measures of organ function (e.g., uremia, hyperammonemia), as these are mediators on the path between delirium and ADRD.

To produce estimates of cumulative incidence curves standardized to the distribution of baseline variables in our target trial population, we include product (interaction) terms for treatment group assignment and time in the pooled logistic regression model^21^. The probabilities from this model are used to estimate the 2-year predicted probability of developing ADRD in each treatment group, and the difference between the curves represents the estimated effect of delirium on ADRD. The 95% CIs for these curves would be computed via nonparametric bootstrapping with 1000 iterations, such that the analysis is conducted in its entirety in each sample.

### Emulation of the target trial: Ascertainment of eligibility, exposure, and outcomes

We mirrored each protocol component as closely as possible, with modifications made as needed to accommodate our use of observational data (Table 1). We emulated the target trial using observational data from the Massachusetts General Brigham (MGB) electronic medical records (EMR) system from patients hospitalized between April 2017 and September 2019.Retrospective analysis of this data was approved by the Institutional Review Board (IRB), which waived the requirement for informed consent.

Eligibility requirements were ascertained from information in the EMR. To select patients who were community-dwelling, we required evidence in the electronic medical record (EMR) of outpatient primary care visits over the prior 12 months. To verify the absence of ADRD at baseline, we required that patients had no ADRD-related medications or diagnostic (ICD) codes in the EMR during this period.

Incident delirium was ascertained by a previously published Natural Language Processing (NLP) algorithm, which was validated against the Confusion Assessment Method (CAM) scores and other markers of delirium^27^. This algorithm analyzes EMR clinical notes each day to determine whether a patient was delirious that day. Unlike International Classification of Diseases (ICD) codes for delirium, which identify delirium in EMR data with high specificity but low sensitivity^28^, the NLP algorithm is both highly sensitive and specific, with accuracy comparable to manual chart review^27^. The NLP algorithm thus makes large-scale delirium studies like the present one feasible. This NLP algorithm was developed on the same population as this study.

The primary outcome, incident ADRD, was ascertained by defining the presence of either an ADRD ICD code (290.X, 294.X, 331.X, 780.93, G30.X, G31.X) or an ADRD medication (Donepezil, Galantamine, Rivastigmine, and their respective brand names Aricept, Razadyne, Exelon) in the patient’s record within the 12 months prior to hospitalization^29^. Time to ADRD was calculated as the number of days between the patient’s admission date and the day evidence of ADRD first appeared in their medical record, which we considered the approximate time of ADRD diagnosis.

Death is a competing risk for the primary outcome, ADRD. We extracted the death date from the EMR demographics data file (Death Master File). Mass General Brigham updates death data monthly from the Social Security Administration. Thus, deaths were captured even if the patient was transferred to a nursing home or another non-Mass General Brigham facility.

### Target trial emulation

#### Emulation of randomization

In the target trial, balanced baseline characteristics would be attained through randomization. In our emulation of the target trial, we emulated randomization by gathering information on clinical and sociodemographic characteristics, assessing differences in their distribution between treated and nontreated groups, and standardizing for relevant confounders in the analysis. In addition, we used cloning (see below) to further emulate randomization. Baseline variables identified as potential sources of confounding to be adjusted for are the same as described above for our target trial (age, sex, race, and burden of chronic illness (CCI)). Cloning is an established, valid approach to emulate randomization^30-32^.

#### Emulation of the grace period

The target trial includes a 14-day grace period from randomization (time zero, t_0_), during which patients randomized to the delirium group must develop delirium (Figure 2). A consequence of having a grace period is that, during it, a patient’s observational data is consistent with more than 1 treatment strategy. For example, in our emulated trial, a patient who has not developed delirium by day 10 after randomization has data consistent with both the “develop delirium before 14 days” and “never develop delirium” strategies. Consequently, we created 2 identical copies (clones) of each individual’s data, assigned each to 1 of the strategies, and censored it when the data became inconsistent with its assigned strategy^33,34^. Note that cloning also has the benefit of balancing baseline covariates between the two treatment groups at trial entry.

**Figure 2.**
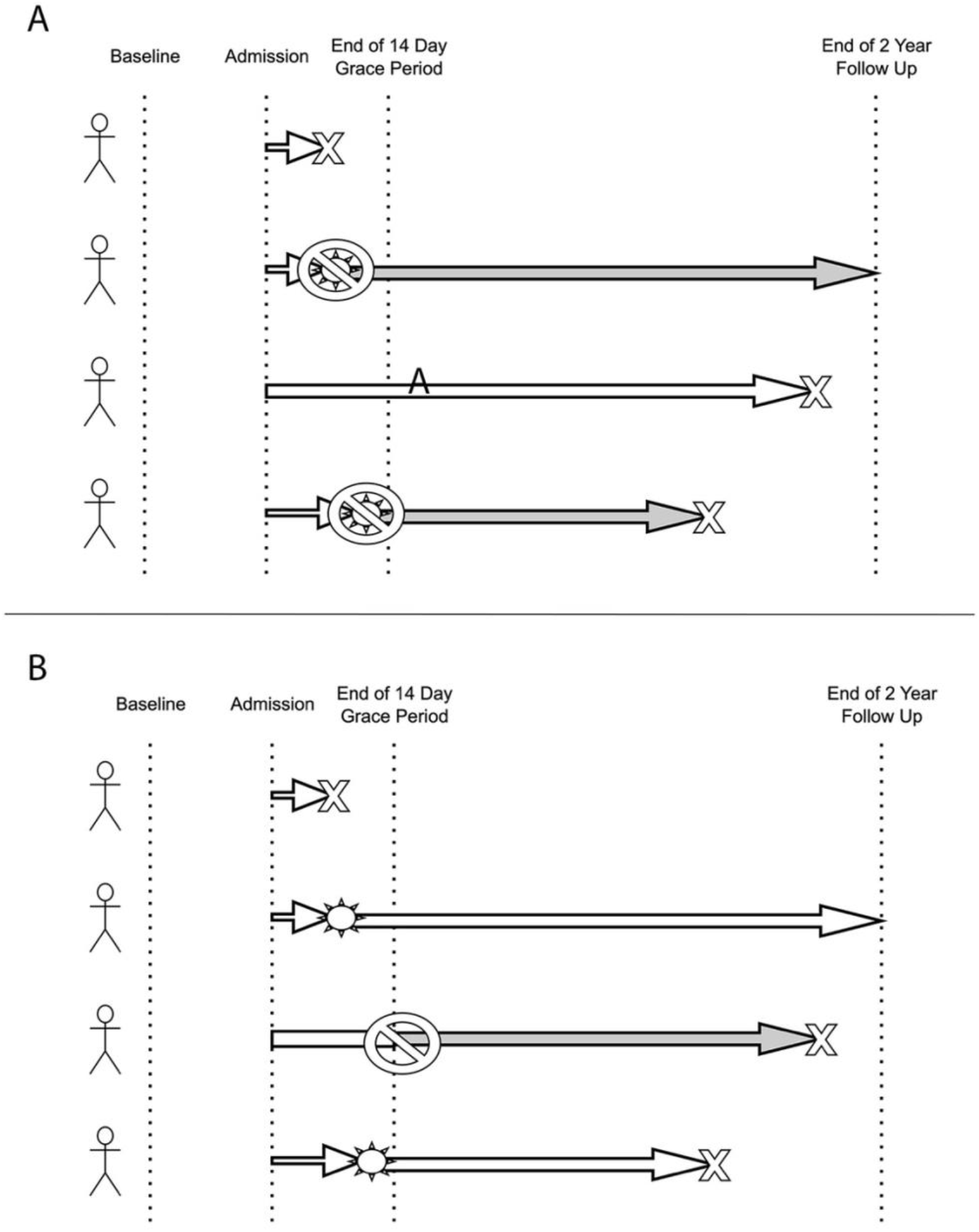
Timeline of censoring in the target trial and our emulation of the target trial. Administrative censoring occurs if an individual does not develop the primary outcome of ADRD during the follow-up period. Non-administrative censoring occurs when patients deviate from the target trial protocol. Grace period censoring occurs during the 14-day grace period: patients in the placebo group are censored if they develop delirium on the day it occurs, and patients in the treatment group are censored at 14 days if they do not develop delirium.Censoring due to death or loss to follow-up occurs during the follow-up period when individuals do not show up for follow-up appointments. **(A)** Censoring diagram for the control group. **(B)** Censoring diagram for the exposure group. Sun Symbol = patient develops delirium, “No” Symbol = patient is censored.

#### Statistical analysis of the emulated trial

We estimated the effects within the emulated trial using pooled logistic regression as we would have for the target trial (see above), using inverse probability weighting or the g-formula to adjust for baseline confounding.

## RESULTS

Among patients admitted within the emulated trial period, 2225 were eligible for our emulated trial. Figure 3 shows the selection process. Table 2 summarizes the cohort’s characteristics. Overall, 51.9% of patients were female; among those who developed delirium, 76.2% were Caucasian; among those who did not develop delirium, 46.0% were female, and 76.9% were Caucasian. 964 (43.3%) of patients developed delirium. The average (unadjusted) time to develop delirium in those who developed it was 2 days. The average (unadjusted) time to the diagnosis of ADRD in those who developed it was 297 days. Patients who developed delirium tended to be older (mean age 78.5 vs 75.1). Among those who developed ADRD, those who experienced delirium also developed ADRD earlier (258 vs 367 days). Note that these differences are raw, unadjusted associations rather than estimates of the association of interest. In the emulated trial, all covariates were balanced at baseline across treatment groups due to cloning.

**Table 2.** Cohort Characteristics.

| Variable | Delirium (n = 964) | No Delirium (n = 1261) | p-value<br>(t-test for continuous value;<br>chi square test for discrete value) |
| --- | --- | --- | --- |
| Age, yr [mean, standard deviation] | 78.5 (8.7) | 75.1 (7.5) | $2.2 \times 10^{-16}$ |
| Female sex (%) | 51.9 | 46.0 | 0.006 |
| White/Caucasian Race (%) | 76.2 | 76.9 | n.s. |
| Time to ADRD (day) | 258 (202) | 367 (185) | 0.016 |
| Charlson Comorbidity Index | 1.6 (1.5) | 1.6 (1.6) | n.s. |
| Admitting Location (Floor/<br>Critical Care) (%) | 98.3/ 1.7 | 98.2/ 1.8 | 0.0009 |
| ADRD (%) | 6.2 | 3.3 | 0.001 |

**Figure 3.**
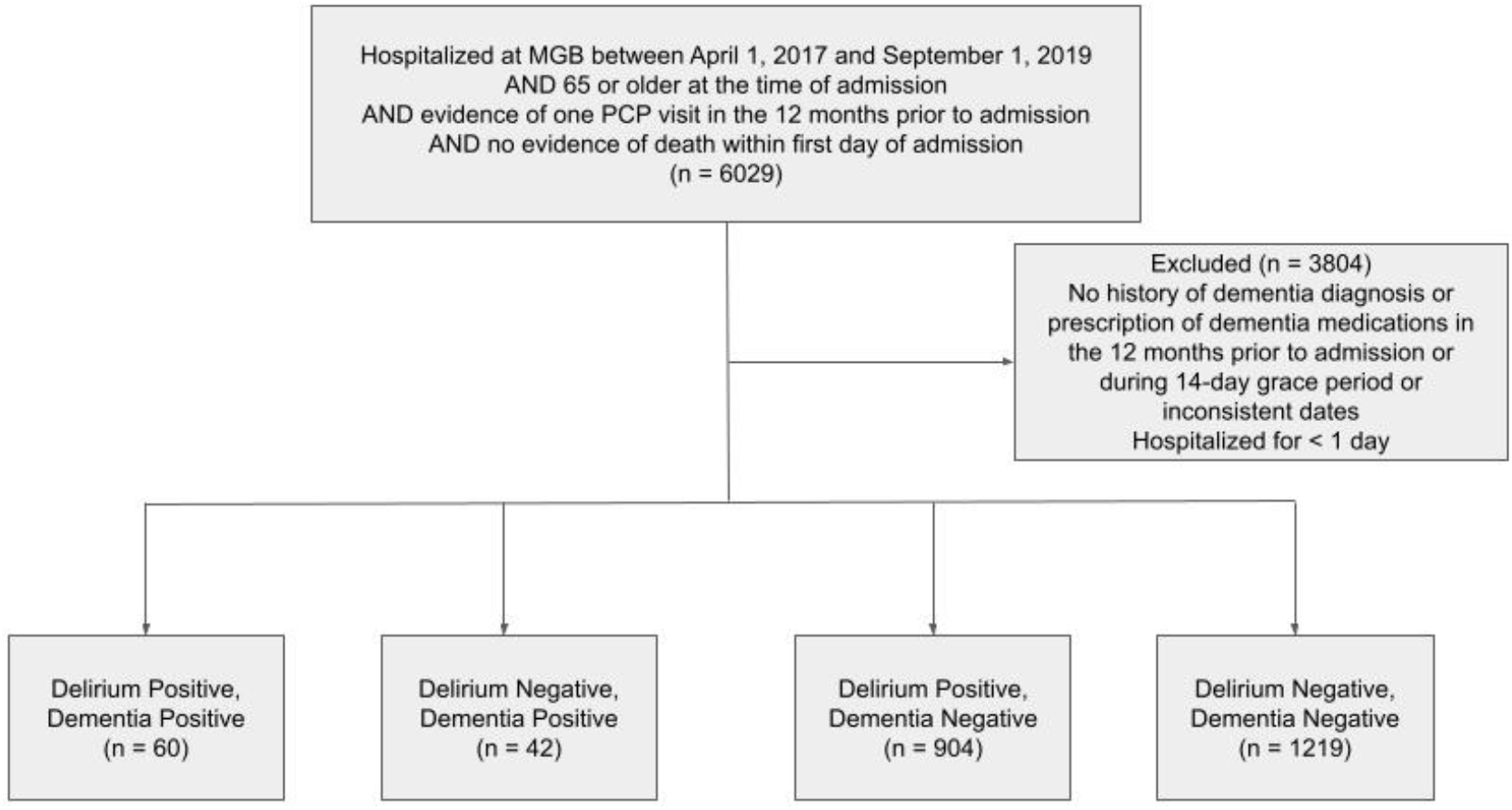
Consort Diagram. 6029 individuals met the initial exclusion criteria. 101 more were excluded due to prior dementia diagnosis. 3 were excluded due to inconsistent dates (death date prior to ADRD or delirium diagnosis date). 24 were excluded due to loss to follow-up, being prior to delirium diagnosis, as they would not contribute information about whether they developed ADRD. The remaining 5901 were included in the analysis.

### Cumulative Incidence

During the 2-year follow-up, there were 352 deaths (15.8% of the cohort), and 83 patients (3.7% of the cohort) developed the primary outcome of ADRD. 75% of patients lost to follow-up occurred in the first 30 days.

At two years post-admission, the adjusted cumulative incidence of ADRD in individuals who did not experience delirium was 7.6% (95% Confidence Interval [CI] 4.0-12.1%) while it was 20.2% (95% CI 13.2-27.9%) for those who did experience delirium when calculated using the IPW method. This was consistent with estimates from the g-formula, with cumulative incidence of ADRD of 9.4% (95% CI 6.3-13.1%) in those who did not experience delirium and 23.3% (95% CI 16.4-31.0%) in those who did. Cumulative incidence curves are shown in Figure 4.

**Figure 4.**
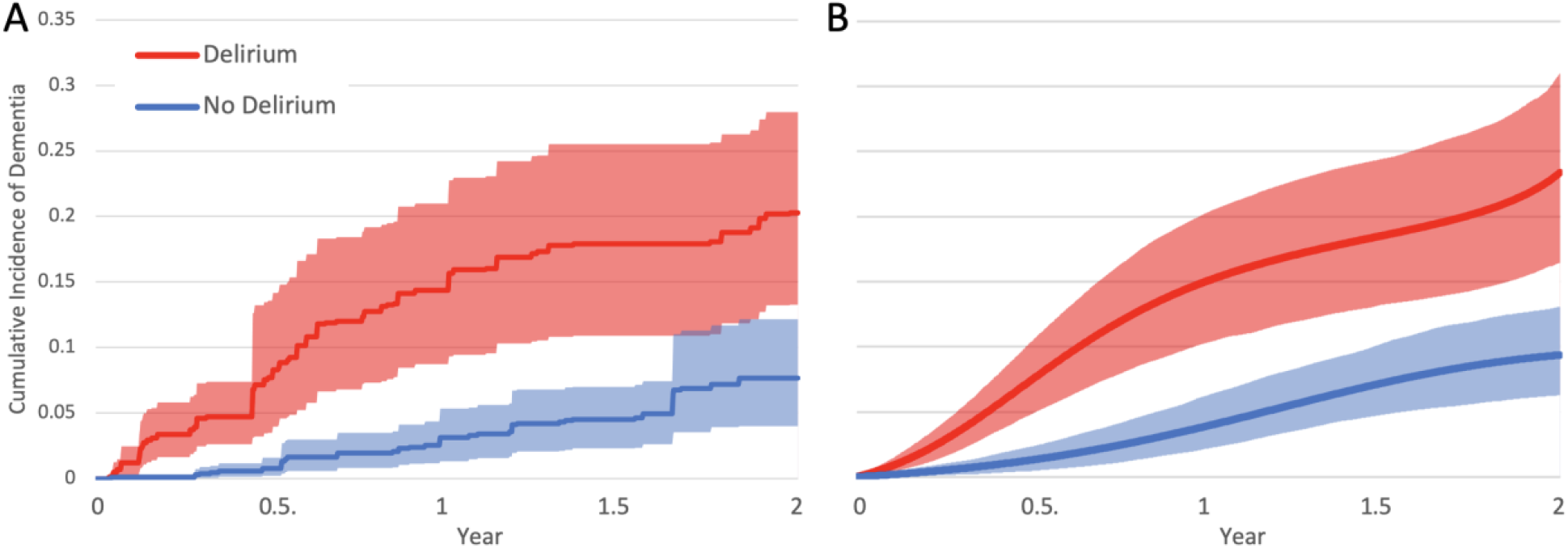
Cumulative Incidence Curves. Cumulative incidence of ADRD in those who experienced delirium versus those who did not. The mean for each exposure group is depicted with a solid line, with the upper and lower bounds depicted by the dashed lines. **(A)** Results using the IPW Method. **(B)** Results using the g-formula method.

### Sensitivity Analyses

To evaluate how features of the analysis affect the results, we conducted two sensitivity analyses. First, we plotted the results without inverse probability weighting, using the Aalen-Johansen estimator (Supplementary Figure 1). This resulted in a cumulative incidence of 1.3% (95% CI 0.8-1.9%) among those with delirium and 2.4% (95% CI 1.7-3.0%) among those without. This demonstrated that not weighting individuals appropriately alters the results. We then evaluated what would occur if the model did not include baseline covariates (Age, Sex, Race, CCI Score) using the g-formula approach (Supplementary Figure 2). This resulted in a cumulative incidence of 1.1% (95% CI 0.7-1.4%) among those with delirium and 2.8% (95% CI 2.2-3.5%) among those without. There is still a significant difference in the two cohorts under these parameters.

We calculated the E-value for sensitivity analysis against unmeasured confounding. At two years post admission, the risk ratio (RR) of developing dementia after experiencing delirium during the hospitalization versus not experiencing delirium is 2.5 (95% CI 2.4 - 2.6) based on g-formula and 2.7 (95% CI 2.3-3.3) based on IPW. The lower bound of the E-value is 4.4 for the g-formula and 4.8 for IPW. To remove the impact of different estimation methods, we use the lower E-value: 4.4, which means that an unmeasured confounder would need to have a minimum risk ratio of 4.4 for both the exposure (delirium) and the outcome (dementia) to render the effect estimate insignificant after adjusting for the unmeasured confounding. While we have adjusted for the major confounders, unmeasured confounders may include genetics, inflammation, and lifestyle factors. Among them, genetics and lifestyle are unlikely to achieve a risk ratio of 3. Inflammation is indeed likely. For example, in a study^35^ of 553 participants above 70 years old, there is a strong relationship between high plasma C-reactive protein (CRP≥3mg/L) and delirium incidence at a risk ratio of 3.0 (95% CI 1.4-6.7) in APOE ε4 carriers, but no association in non-ε4 carriers. Therefore, given the approximately 25% prevalence of APOE ε4 carriers, the overall risk ratio for CRP and delirium is about 1.5, consistent with another study^36^. On the other hand, there are also studies demonstrating a significant association between CRP and dementia. For example, in a study^37^ of 111,242 adult participants, the hazard ratio is 1.69 (95% CI 1.29-2.16) when comparing low plasma CRP to high based on percentiles. In another study^38^ of 4,601 participants above 50 years old, those with mixed Alzheimer’s disease had an odds ratio of 2.14 (95% CI 1.19-3.85) for having high CRP. Although these associations are not on the risk ratio scale, they are unlikely to reach a risk ratio of 3.0. In summary, our effect estimate is robust to unmeasured confounding.

## DISCUSSION

Our emulated trial results suggest that developing delirium during hospitalization more than doubles the risk of developing ADRD in the two years following admission in individuals 65 and older. If our emulated trial design is valid, this result can be interpreted as a strong association between delirium and ADRD. The result is consistent with several prior association studies ^1-6,39-43^, but goes beyond prior findings by emulating a clinical trial and showing a strong relationship between delirium and ADRD.

The possibility of a link between delirium and ADRD has biological support, based on recent evidence suggesting that delirium increases neuroinflammatory and neuronal injury/damage, which potentially contributes to cognitive decline^44^. Biomarker studies have underscored the role of these factors, along with systemic inflammation, in the pathophysiology of delirium^23^. These factors highlight the potential path between delirium and dementia; delirium could cause this damage and thus lead to the accelerated onset of dementia. Our analysis is consistent with the association between delirium and ADRD demonstrated in multiple studie^2,41,43^.

The implications of demonstrating this relationship underscore the importance of delirium-mitigating interventions for long-term cognitive outcomes. Well-supported multicomponent non-pharmacologic interventions for delirium prevention, such as the Hospital Elder Life Program (HELP), can affect not only the incidence of delirium but also the speed of onset of cognitive decline^11^. HELP is a delirium-mitigating technique that utilizes interventions such as ambulation, ensuring patients have hearing aids if needed, etc. Dexmedetomidine has also shown promise in mitigating delirium in hospitalized individuals^14^.

This study has important limitations. We attempted to emulate a target randomized controlled trial; thus, the drawbacks of this approach include the assumptions made when using observational data. These assumptions include the assumption that if a patient had a record of a primary care visit in the year prior to hospitalization, we can be sufficiently confident that, if the patient had pre-existing ADRD, it would have been detected, allowing us to exclude them from our study. The inclusion of only those with PCP visits in the past year was also to ensure that they would be likely to continue to follow up with this provider during the two-year follow-up period, so that the incidence of ADRD would be reflected in the medical record. These assumptions are not perfect, as there could be individuals suffering from ADRD without an ICD code or ADRD medication in their medical records. There is also a lag in the diagnosis of ADRD after symptoms begin, so this follow-up period of 2 years may not have captured many of those who were diagnosed later. Nevertheless, a strength of our study is that all assumptions made in creating our emulated trial are explicit.

We are also limited by the extent to which delirium can be clearly defined. There is a broad range of severity and the possibility of it going undetected, especially in its hypoactive forms. Under-detection may lead this analysis to underestimate the association. Delving further into the severity of delirium and how that affects outcomes could be a worthwhile future investigation.

There are also possible limitations in not controlling for all confounders that could have been addressed with randomization in a controlled trial. This study leverages EMR, so we do not definitively know the baseline cognitive status of the eligible sample. There is the chance of undiagnosed ADRD and thereafter having a diagnosis detected due to the use of the system after hospitalization, not because of delirium. We would expect an increase in ADRD diagnoses early in our follow-up period if this were the case, though this was not observed in our results. By extension, we are also limited to the data in this EMR, without access to other registries. This was seen in the significant lack of race in a portion of this patient population. While we attempted to control for relevant baseline variables, it is not possible to fully rule out unmeasured confounding. Additional variables to consider in the future could include baseline medication use, access to the EMR prior to hospitalization, reason for admission, language, geriatric depression scale, baseline Mini-Mental State Examination, Rey Auditory Verbal Learning Test, and evidence of a brain lesion on MRI. In addition, and importantly, another limitation is that using EMR data does not reflect incidence in the general population. This cohort is limited in diversity, which reduces generalizability and underscores the need for further studies with a more diverse population.

In summary, this study provides evidence of a strong relationship between delirium and ADRD. This relationship should be explored further with interventional trials that use delirium- prevention strategies to reduce the long-term incidence of ADRD following hospitalization in older adults.

## DATA AVAILABILITY

All data produced in the present study are available upon reasonable request to the authors.

## SUPPLEMENTARY MATERIALS

**Supplemental Figure 1.**
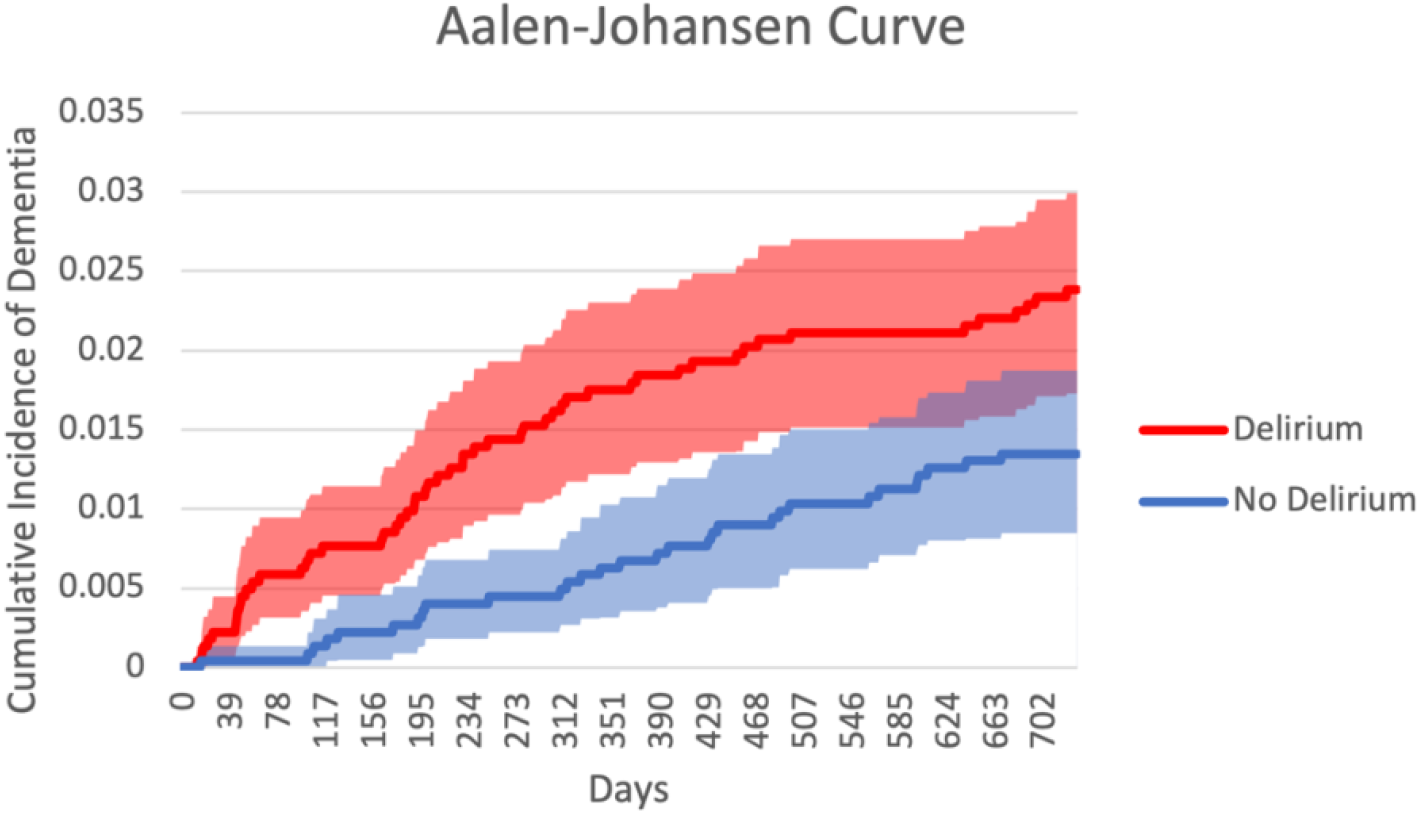
Sensitivity Analysis 1. The effect of not weighting, which is the Aalen-Johansen Estimator. The mean for each exposure group is depicted with a solid line, with the upper and lower bounds depicted by the dashed lines.

**Supplemental Figure 2.**
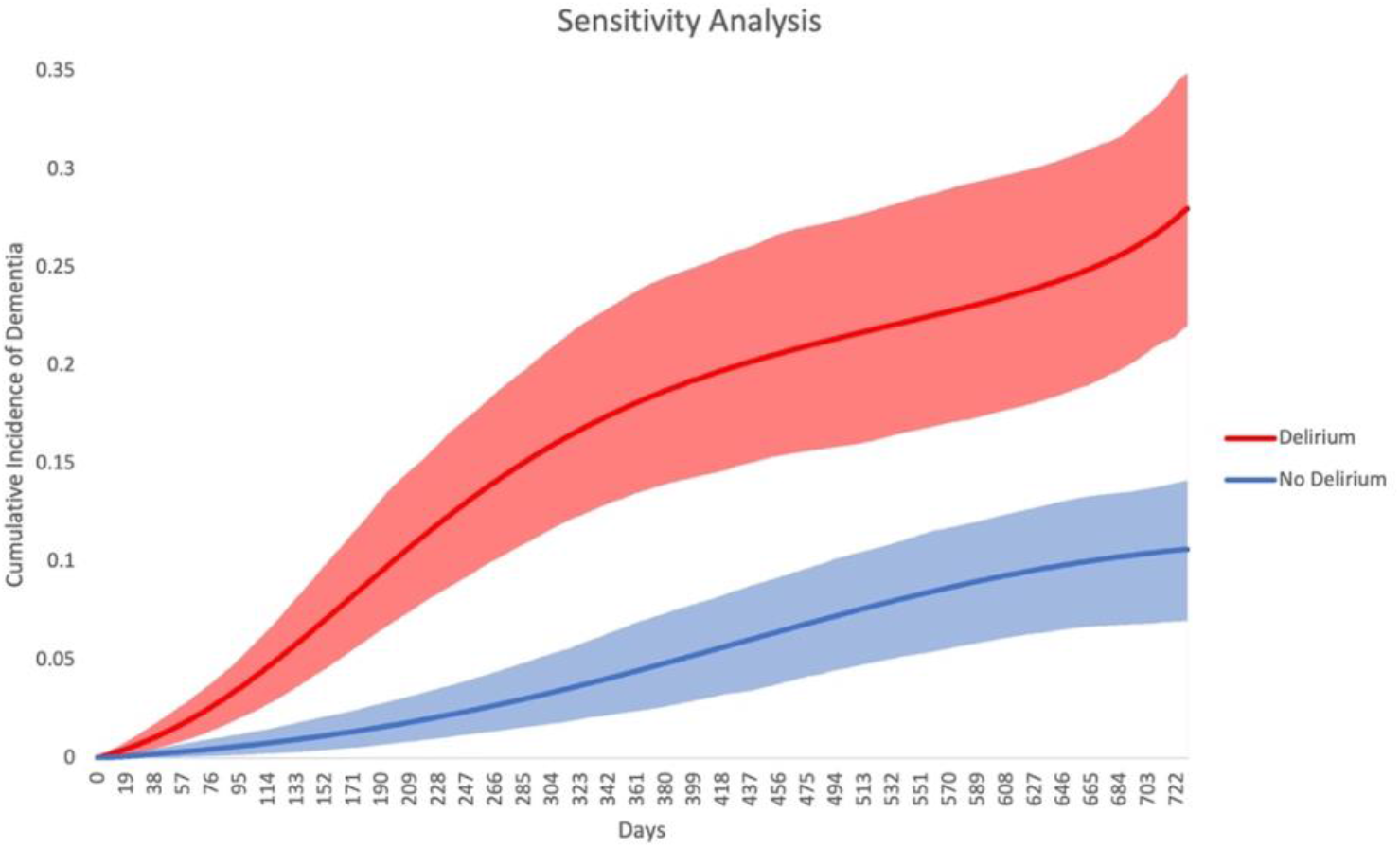
Sensitivity Analysis 2. The effect of not including covariates while using the g-formula method. The mean for each exposure group is depicted with a solid line, with the upper and lower bound depicted by the dashed lines.

